# Multisite longitudinal efficacy trial of a disclosure intervention (TRACK) for HIV+ mothers: An update

**DOI:** 10.1101/2022.08.26.22278479

**Authors:** Debra A. Murphy, William D. Marelich, Lisa Armistead, Marya T. Schulte

**Affiliations:** Semel Institute for Neuroscience and Human Behavior, University of California, Los Angeles; Department of Psychology, California State University, Fullerton; Department of Psychology, Georgia State University

## Abstract

**Background:** The <u>T</u>eaching, <u>R</u>aising, <u>A</u>nd <u>C</u>ommunicating with <u>Ki</u>ds (TRACK) intervention full-scale longitudinal efficacy trial was published in 2021 and was based on a previously successful pilot intervention published in 2011. The TRACK intervention assists mothers living with HIV (MLH) with serostatus disclosure to their young children. In the publication reporting the full-scale trial, TRACK MLH were four times more likely to disclose their HIV serostatus than controls, with the rate increasing to six times more likely applying GEE logistic regression. Intervention MLH showed improvements in communication, social support, family routines, and disclosure self-efficacy; they also demonstrated decreased anxiety and better physical and mental health scores; and their children reported significantly more decline in worry than controls.

**Methods:** This preprint presents recent analyses showing increases in the reported HIV disclosure rates over and above those reported in the full-scale trial, due to additional follow-up datapoints received after the 2021 publication.

**Results:** For MLH in the intervention group, disclosure is 37.6% compared to 34.1% from the 2021 publication, and 12.1% in the control condition compared to 11%. Odds of disclosing if participants were in the intervention group are 4.39 (95% CI. = 2.0, 9.5) compared to 4.20 (95% CI. = 1.8, 9.3), with the intervention group 30% more likely to disclose compared to the 28% reported in 2021. Regarding site disclosure rates for the intervention group, the California disclosure rate is 43.6% compared to 35.9% previously reported; Georgia’s site rate remained unchanged.

**Conclusions:** These updates are now included in our final analysis dataset and show the intervention was more successful than previously reported.

## Introduction

The <u>T</u>eaching, <u>R</u>aising, <u>A</u>nd <u>C</u>ommunicating with <u>Ki</u>ds (TRACK) intervention was developed from both longitudinal quantitative and qualitative work of the first [1,2,3] and third [4] authors, utilizing an integrative model of disclosure [5]. The three-session intervention aimed to equip MLH with skills and confidence to complete disclosure with their well children by impacting relationship context variables targeted for intervention within the integrative model and building MLHs’ self-efficacy for disclosure. Initially, a pilot study of the TRACK intervention [6] successfully increased mothers’ disclosure to their young children (6 – 12 years), with the intervention group six times more likely to disclose. Following the pilot study, some revisions were made to the TRACK intervention to strengthen it based on the pilot trial results and feedback from the participants. A full-scale longitudinal efficacy trial [7] was conducted that included behavioral exercises within three individual sessions, with assessments conducted at baseline, 3-, 9-, and 15-month follow-ups. The intervention specifically targeted intermediate variables of communication, parental coping, family routines, disclosure self-efficacy, and secondary outcome variables for MLH’s physical and mental health, children’s mental health, and family relationships/functioning.

In this study, we found increases in our reported HIV disclosure rates from the 2021 full scale trial publication, due to additional follow-up datapoints received after publication. The updates were received in December 2021 after publication of the 2021 article, consisting of 23 datapoints from 20 study participants recruited later in the study protocol (the majority received were for the 15-month follow-up period). These updates are now included in our final analysis dataset and show the intervention was more successful than previously reported. Although the addition of these data yielded no changes of practical significance to the 2021 article, we are providing this update to provide the updated disclosure rate, and for future replication purposes as our analysis datasets will be made publicly available to researchers.

## Method

The methodological approach used in this report is the same as that described in detail previously [7]. The intervention key components are also described in detail in the TRACK implementation paper [8]. Briefly, MLH (mean age = 39.27, SD 7.89) and their children (6-14 years) were recruited at two sites (Southern California and Atlanta) and randomized to an intervention or wait-list control group (N = 176 dyads). The TRACK intervention includes behavioral exercises within three individual sessions; assessments were conducted at baseline, 3-, 9-, and 15-month follow-ups. A conceptual model based on integrative disclosure theory [5] was utilized; this model hypothesized that key variables targeted in the intervention—disclosure self-efficacy, parent-child communication, and family routines—could lead to a choice of disclosure by MLHs, which in turn would lead to improved maternal and child mental health and improvements in the parent-child relationship. The overall analysis design was a randomized pretest–posttest two-group design with repeated assessments.

Measures included the following constructs: demographics, family communication, parenting skills (including family routines), disclosure or non-disclosure (including self-efficacy for disclosure), maternal physical and mental health, child mental health, and family functioning.

All study procedures were approved by the appropriate Institutional Review Boards at both sites. This study has been registered with ClinicalTrails.gov with registration #NCT01922206.

## Results

For our full sample of mothers living with HIV (MLH), disclosure is 24.4% (43 of 176) compared to 22% reported in 2021 paper. For MLH in the intervention group, disclosure is 37.6% (32 of 85) compared to 34.1% from 2021, and 12.1% (11 of 91) in the control condition compared to 11%. Odds of disclosing if participants were in the intervention group are 4.39 (95% CI. = 2.0, 9.5) compared to 4.20 (95% CI. = 1.8, 9.3) previously noted, with the intervention group 30% more likely to disclose compared to 28% reported in 2021. Regarding site disclosure rates for those in the intervention group, the California disclosure rate is 43.6% compared to 35.9% previously reported (the 7.7% increase for California is not significant based on test of proportions); Georgia’s site rate remained unchanged. Rate changes in disclosure over time for the intervention group are: 15.3% at 3-month follow-up (unchanged), 12.9% at 9-month follow-up (10.6% reported in 2021), and 9.4% at 15-month follow-up (8.2% prior). For the control condition, the only change was at 15-month follow-up (5.5%; 4.4% previously reported).

Mixed-model repeated measures mean estimates, standard errors, and pairwise comparisons noted in Tables 1 and 2 of the 2021 article remain fundamentally unchanged (due to a production issue, some Table 2 degrees of freedom values were assigned negative signs – these should be encased in parentheses). We adhered to a strict alpha level (*p* </= .05) to evaluate mean differences within groups across time, and a few secondary findings are now *p* </= .07 but did not yield changes in effect sizes, and for all but one finding, other time-based comparisons continue to support differences at *p* </= .05.

Only a few secondary findings changed, but do not any impact on the main conclusions of the 2021 paper or the Discussion. These are as follows. *Intervention group*: Two findings for MLH no longer meet the *p* </= .05 cutoff (MOS general health [*p* = .06] and MOS vitality [*p* = .07]), and two findings for child outcomes no longer meet *p* </= .05 (CDI Total score baseline to 3-month FU [*p* = .06], and IPPA Lack of Alienation baseline to 3-month FU [*p* = .07]). *Control condition*: Two findings for child outcomes no longer meet the *p* </= .05 cutoff (CDI Ineffectiveness Baseline to 15-mo FU, *p* = .06; IPPA Communication, Baseline to 3-mo FU, *p* = .06). One finding for the child outcome Worry (Baseline to 3-mo FU) now meets the *p* </= .05 cutoff at *p* = .04. However, as noted, the primary outcomes of the paper remain as reported: (a) that the intervention increased disclosure significantly; and (b) that the targeted intermediate variables hypothesized as necessary for efficacious disclosure all remain significant (disclosure self-efficacy, parent-child communication, parental coping, and family routines).

## Discussion

The TRACK intervention was even more successful than indicated in the full-scale trial article [7]. Due to additional follow-up datapoints received after the 2021 publication--primarily from the 15-month follow-up--we are able in this preprint to show improved disclosure rates while confirming that all the components that we originally hypothesized to be necessary for an efficacious disclosure as early as the pilot study are critical to include in the intervention. Overall, for MLH in the intervention group the disclosure rate is 37.6%: the California disclosure rate is 43.6% and the Georgia rate is 32.6%. In addition to the strength of the TRACK intervention to promote maternal disclosure of HIV, improvements were also found in family cohesion, stability of family routines, and maternal well-being. Thus, TRACK has implications for family interventions in general, not just families who are affected by HIV/AIDS.

It should be noted that disclosure was defined in this study as full disclosure—that is, the MLH had to disclose that she was HIV-infected; partial disclosures such as saying she was ill or had a virus, without specifying HIV, were not counted as a disclosure. Moreover, the intervention was designed to mitigate any anxiety or depression the child may feel upon being made aware of their mother’s serostatus. The MLH practiced behavioral skills aimed at improving communication with their children about the illness in a developmentally appropriate way while limiting the child’s distress. In summary, these were full disclosures and per MLH report they were done appropriately (e.g., keeping an appropriate emotional tone during disclosure, being prepared to answer questions about HIV transmission, waiting to disclose until being in the right frame of mind). These new findings strongly indicate that the TRACK intervention can change mothers’ behavior to affect disclosure of her HIV status to her child(ren) as well as provide the skills to do so in a way that improves mental health within the family.

## Supporting information

CONSORT

## Data Availability

All data produced in the present study are available upon reasonable request to the authors.

## Authors’ contributions

DAM and LA conceptualized and designed the study. WDM conducted all analyses and prepared the results section. DAM wrote the Introduction, Method, and Discussion. MTS reviewed and edited the draft. All authors provided valuable feedback on the implications of the study and contributed to and approved the final manuscript.

## Acknowledgements

We would like to thank the TRACK participants—both mothers living with HIV/AIDS and their children—who participated in the study.

