## Supplementary material for "Multisite longitudinal efficacy trial of a disclosure intervention (TRACK) for HIV+ mothers: An update": CONSORT

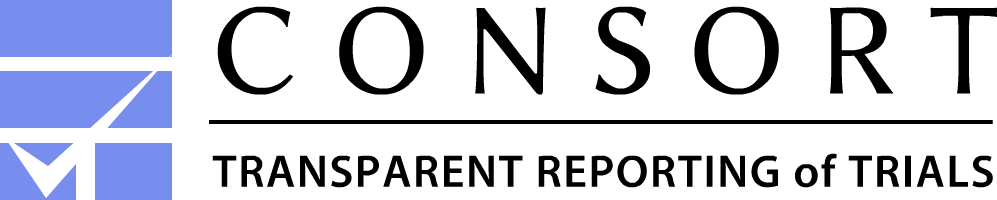

**CONSORT Appelbaum et al. 2018 Adaptation Flow Diagram**

**Assignment**

**Analysis**

**Follow-Up**

**Enrollment**

Assessed for eligibility (n= 262)

Excluded (total n= 86) because

♦  Did not meet inclusion criteria (n= 76)

♦  Refused to participate (n= 0)

♦  Other reasons (n=10) removed after baseline because deemed not to meet inclusion criteria after first assessment

Analyzed (n= 85)
♦ Excluded from analysis (n= 0) Reasons:

Lost to follow-up (n= 0) Reasons:

Discontinued participation (n= 0) Reasons:

Assigned to TRACK intervention (n= 85)

♦ Received TRACK intervention (n= 80)

♦ Did not receive TRACK intervention (n= 5) Reasons: moved outside of state, could not locate, refused to participate

Lost to follow-up (n= 0) Reasons:

Discontinued participation (n= 0) Reasons:

Assigned to Wait-list Control group (n= 91)

♦ Received Wait-List intervention (n= 39)

♦ Did not receive Wait-List intervention (n= 52) Reasons: unable to attend, not interested at that point in time

Analyzed (n= 91)
♦ Excluded from analysis (n= 0) Reasons:
